# Large Language Models for Detecting Body Weight Changes as Side Effects of Antidepressants in User-Generated Online Content

**DOI:** 10.1101/2023.12.09.23299754

**Authors:** Taïoh Yokoyama, Johan Natter, Julien Godet

## Abstract

**Objective:** Healthcare websites allow patients to share their experiences with their treatments. Drug testimonials provide useful information for real-world evidence, particularly on the occurrence of side effects that may be underreported. We investigated the potential of large language models (LLMs) for detecting signals of body weight change as under-reported side effect of antidepressants in user-generated online content.

**Materials and Methods:** A database of 8,000 user-generated comments about the 32 FDA-approved antidepressants was collected from healthcare social websites. These comments were manually annotated under the supervision of drug experts. Several pre-trained LLMs derived from BERT were fine-tuned to automatically classify comments describing weight gain, weight loss, or the absence of reference to a weight change. Zero-shot classification was also performed. Performance was evaluated on a test set by measuring the weighted precision, recall, F1-score and the prediction accuracy.

**Results:** After fine-tuning, most of the BERT-derived LLMs showed weighted F1-scores above 97%. LLMs with higher number of parameters used in zero-shot classification almost reached the same performance. The main source of errors in predictions came from situations where the machine predicted falsely weight gain or loss, because the text mentioned these elements but for a different molecule than the one for which the comment was written.

**Conclusion:** Even fine-tuned LLMs with limited numbers of parameters showed interesting results for the detection of adverse events from online patient testimonials, suggesting they can be used at scale for real-world evidence.

## Introduction

Body weight changes are described adverse effects (AEs) of antidepressant use. Weight loss or gain is also a component of the clinical presentation of depression. Consequently, a change in body weight can be considered a potentially expected outcome in treated patients and weight changes may not always be readily attributed to the use of these treatments as adverse effects. However, if they do occur, they can significantly impact patients and their therapeutic management, as these changes directly affect self-image and adherence to treatment. AEs like weight changes become more evident with long term treatment. They are thus difficult to capture in clinical trials due to their short follow-up period (1). Their identification and correct quantification rely on further evaluation in real-world conditions, mostly through clinical reporting to pharmacovigilance. But attributing to treatments long-term and delayed changes is more difficult than evidencing acute immediate effects.

The first-generation of antidepressants are known to cause weight gain (2). These effects can also be observed with more recent treatments (3). It has been reported that the general trend over 10 years for people who have received an antidepressant treatment is weight gain (4). The results of cohort studies and meta-analyses also agree to identify some antidepressants being at higher risk (1). Molecules such as mirtazapine, amitriptyline, and paroxetine are generally associated with weight gain, while bupropion and fluoxetine seem to be more associated with weight loss (1). However, for many molecules, both weight gain and weight loss are reported, highlighting the variability in individual responses to these medications as well as the importance of temporal aspects, with effects that may differ at the start of treatment or over the long term. Owing to all these elements, patient testimonials can be a very interesting source of information to better describe the adverse effects of this family of drugs.

The democratization of the internet has led to the emergence of many platforms for free expression, including online health communities. These health-specialized sites allow patients to share their experiences with their treatment. These testimonials are a potential source of massive data for collecting information about the effects of treatments in real-world conditions (5). The diversity of information available in these testimonials is a direct result of a proactive and patient-centered information sharing approach (6**? ?**). It is conceivable that the adverse events with the most significant impact on patients are the most commented on, even if they may not be the ones that are discussed the most between patients and caregivers during medical consultations and therefore not the ones that are reported the most. This source of information is an interesting alternative in the context of adverse events that would be under-reported in traditional pharmacovigilance channels (7, 8). The main disadvantages of these data are that they are unstructured, with a wide linguistic diversity, which complicates the automation of their analysis. Given the number of comments generated, processing by human operators is proving to be a long and tedious task. During the 2010’s, natural language processing (NLP) tools (9) coupled to deep neural networks have shown promising results for AEs detection in text data (10), and a variety of architectures have been proposed to detect AE in text sources (11–13). Automated AEs were screened scrutinizing electronic health records of patients (14–17), and Web data including social networks like X (formerly Twitter) (6, 18–22). Most of these works aimed at identifying the mention to an AEs and did not focus specifically on the detection of a particular AE.

**Fig. 1.**
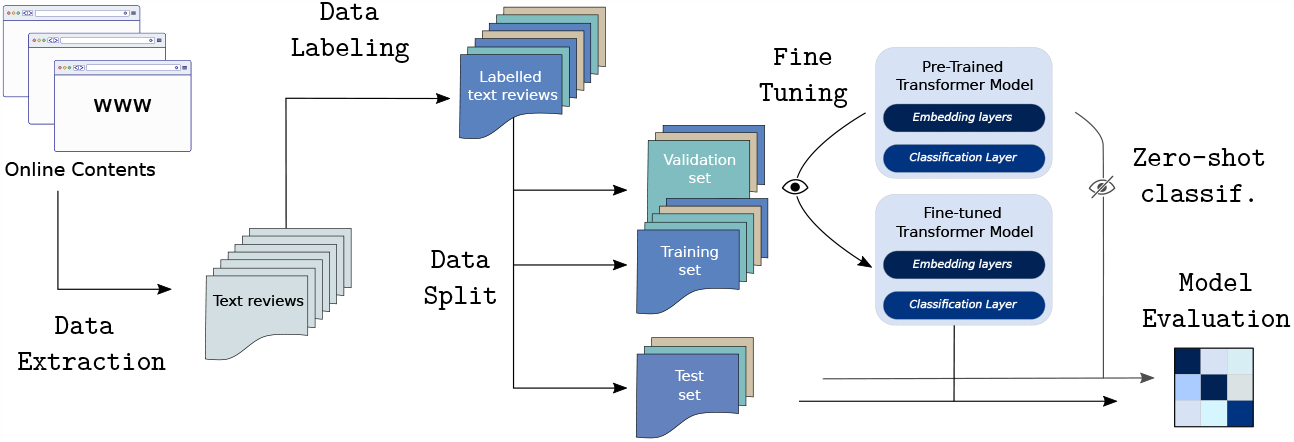
Data collection, Transfer learning or Zero-shot classification approach to text classification. BERT pre-trained transformers are key elements. Models have been fine-tuned using a training and a validation set of labeled text reviews - or used *as is* for zero-shot classification. Classification performance was performed on a test set.

More recently, the development of transformer-based models has revolutionized NLP and have become the new standard for many NLP tasks (23). Few recently published works make use of pretrained transformer-based models for ADE extraction on informal texts (24), especially the models based on pre-trained models like bidirectional encoder representations from transformers (BERT) (21, 25–29). Self-attention (or QKV-attention) is central mechanism in Transfomer -allowing the model to attend to different parts of the text input sequence when making predictions and to learn long-range dependencies in the input sequence. Transformer models can be trained on large datasets to learn language representations. Pre-trained models can then be used for zero-shot classification or they can be fine-tuned for a specific classification task. In zero-shot classification, pre-trained models are used without any specific additional training examples for new classes (30). Their performance rely on their ability to generalize classification properties based on the language representation they captured during pre-training. On the opposite, the fine-tuning step consists of training further a pre-trained model on a new dataset and for new classification classes to improve the model’s performance on this specific task (31).

Here, we explored the potential of different models of classification to automatically recognize comment texts describing weight gain, weight loss, or the absence of reference to a weight change. In particular, we focused on BERT and BERT-related transformers, that have achieved state-of-the-art results on a wide range of NLP tasks (32). The novelty of BERT was to encode the context of a word from both the left and the right (bidirectional). BERT was designed to pretrain deep bidirectional representations from unlabeled text by jointly conditioning on both left and right context in all layers.

We also used RoBERTa - a robust re-implementation of BERT with some modifications to the key hyperparameters and minor embedding tweaks. It has been shown to outperform BERT on a variety of NLP tasks (33). Roberta uses a byte-pair encoding tokenizer instead of a word-piece tokenizer to better handle rare words and out-of-vocabulary words. It trains with a larger mini-batch size and for a longer number of steps to better learn long-range dependencies in text and it removes the next sentence prediction (NSP) objective, found to be less effective than the masked language modeling (MLM) objective for pre-training BERT models. If BERT and RoBERTa are highly efficient, it can be difficult to run these large models on edge devices or to train or use them with limited computational ressources.

DistilBERT is a distilled version of BERT that is smaller and faster (34). It is trained using a knowledge distillation procedure, which involves training a smaller model to mimic the predictions of a larger model and it has been shown to achieve comparable performance to BERT and RoBERTa. DistilBERT is a good choice for applications where speed, efficiency, and ressources are important. Similarly, we explored SqueezeBERT, a lightweight and efficient transformer-based language model that is specifically designed for mobile and embedded devices (35). SqueezeBERT is based on the BERT architecture, but it has a number of modifications that make it smaller and faster including using grouped convolutions instead of fully-connected layers to reduce the number of parameters in the model.

BERT and these BERT-derived models are pre-trained on a massive dataset of text and code. They can be used pre-trained and only fine-tuned on a specific downstream task including classification without having to be trained from scratch (36). We also explored the performance of DistilBERT and DeBERTa in the context of zero-shot classification. DeBERTa uses disentangled attention for learning richer contextual representations of words and an enhanced mask decoder for more accurate predictions of masked words, providing this model state-of-the-art performance on a variety of natural language processing tasks (37).

Taken together, we aimed to explore the potential of large language models (LLMs) in detecting signals of changes in body weight as potential side effects of antidepressants in user-generated online content. Our investigation included fine-tuning and zero-shot classification using various BERT-based models. Our main motivation was to explore the ability of these models in identifying signals within large datasets of user reviews. Additionally, we aimed to investigate their capacity to distinguish weight gain from weight loss - two closely related elements sharing similar syntax that require a more nuanced analysis and interpretation of the language used in the comments for accurate classification. This represents an additional step beyond signal detection, moving towards assessing the causality of a drug in the occurrence of adverse effects. Most of these different models performed particularly well in this task.

## Material and Methods

### Data collection

Data collection took place between August 2022 and November 2022. The data corresponded to publicly available user-generated online content from four specialized websites at the time of data collection (38): *Drugs*.*com, WebMD, Everyday Health*, and *Ask a Patient. Drugs*.*com, WebMD* and *Everyday Health* are websites that provide information about medications and health to the general public. *Ask a Patient* is a platform where patients can share their first-hand experiences with their prescribed drugs. Data from *Drugs*.*com, WebMD* and *Everyday Health* were scraped using R version 4.2.2 and the R packages rvest (39), httr (40), and xml2 (41) while data from *Ask a Patient* (and the *psyTAR*) database were kindly provided by *Askapatient*.*com*. A random sample of 8,000 texts, stratified by site and drugs, was extracted for manual annotation.

### Data labeling

The annotation was performed by ten operators, including two pharmacists, a general practitioner, two pharmacy students and five non-drug experts. The data set was randomly split in 10 sets with overlap to allow for the measurement of inter-rater agreement. The drug on which the users were commenting on in the input text was identifiable to the rater. Each input text was categorized into three categories: weight gain, weight loss, or the absence of reference to a weight change. However, the label had to be applied on the drug a user was commenting on (i.e. if weight change elements were described only for another drug mentioned in the user-content, like previous experience in treatment history, then the label was set to absence of reference to a weight change for this given drug). In cases where the rater felt uncertain about the appropriate class assignment, he/she could add a specific code to allow this input text be reviewied by a drug-expert.

### Data split

The data set was split into training (n = 6,000), validation (n = 1,000), and test (n = 1,000) datasets. The training and validation data sets were used to adjust the LLMs during the fine-tuning step. The test set was used to evaluate the performance of the model. It is important to note that the training, the validation and the test sets were defined identically for all models in order to facilitate comparisons.

### Large Language Models

The pre-trained models were loaded by instanciating a given LLM configuration model using the generic sequence classification model class available in the Hugging Face library https://huggingface.co/ (42) for Python 3.8.

The different models for zero shot classification were :

- distilbert-base-uncased (6-layer, 768-hidden, 12-heads, 66 M parameters)
- deberta-v3-base (12 layer, 768-hidden,12-heads, 86 M parameters)
- deberta-v3-large (24 layer, 1024-hidden,16-heads, 304 M parameters)

and for fine-tuning :

- bert-base-uncase (12-layer, 768-hidden, 12-heads, 110 M parameters)
- roberta-base (12-layer, 768-hidden, 12-heads, 125 M parameters)
- distilbert-base-uncased (6-layer, 768-hidden, 12-heads, 66 M parameters)
- squeezebert-uncased (12-layer, 768-hidden, 12-heads, 51 M parameters)

The input texts corresponding to user reviews were lower-cased and tokenized using the appropriate pre-trained tokenizer using the AutoTokenizer class from the Transformers model library, without any additional preprocessing steps. In all cases, the tokenized texts were paded and truncated to a fixed length of 512.

The models were trained over 5 epochs, using the AdamW optimizer (43) and a learning rate of 2 *×*10^*−*5^ with batch sizes of 4 to 16, depending on models. Accuracy and F1-score were monitored during training, as well as the training and validation loss. Models were trained for about 30-45 minutes with NVidia 1080 GPU acceleration.

### Prediction performance

The models were evaluated using accuracy, weighted precision, recall and F1-score and ‘macro’ F1-score measures. Performance was measured on the predictions of the test dataset, which was unseen during the training step.

## Results

### Characteristics of the training data

The database included 80,594 comments about 32 different antidepressants, from four websites: Drugs.com (35.8 % [28,876]), WebMD (43.1 % [34,748]), Everyday Health (18.2 % [14,672]), and Ask a Patient (2.9 % [2,298]). Of these comments, 8,000 were randomly selected for manual annotation, stratified by website and antidepressant molecules. The annotation process was designed to allow for some overlap between text inputs so that inter-rater reliability could be estimated. Out of the 8,000 text inputs, 3,500 were evaluated twice, resulting in 96 label discrepancies (2.74 %) reconciled by an expert reviewer. This gave a Krippendorff’s alpha of 0.841. The distribution of labels in the dataset are presented as a function of drugs in Table S1. Following the data split, the training and validation sets comprised 6,000 and 1,000 labeled inputs, respectively, while 1,000 inputs were kept for testing. As expected, labels associated with gaining or losing weight were in the minority in the training and testing sets-with almost 90% of the labels corresponding to an absence of mention of change in body weight in the input text. This led to an accuracy value of 0.897 in the validation dataset for a dummy classifier predicting the most represented class (see Table 1).

**Table 1.** Performance of the different BERT-based LLMs evaluated on a validation dataset of 1,000 text inputs.

|  | Model | Accuracy | Precision<br>weighted | Recall<br>weighted | F1-score<br>weighted | F1-score<br>macro |
| --- | --- | --- | --- | --- | --- | --- |
| Baseline | Dummy classifier | 0.897 | 0.805 | 0.897 | 0.848 | 0.315 |
| Zero-shot | DistilBERT | 0.265 | 0.817 | 0.265 | 0.350 | 0.384 |
|  | DeBERTa *base* | 0.962 | 0.963 | 0.962 | 0.962 | 0.833 |
|  | DeBERTa *large* | **0.964** | **0.971** | **0.964** | **0.966** | **0.869** |
| Fine-tuned | BERT | **0.975** | **0.977** | **0.975** | **0.976** | **0.895** |
|  | RoBERTa | **0.975** | 0.976 | **0.975** | **0.976** | 0.891 |
|  | DistilBERT | 0.973 | 0.975 | 0.973 | 0.974 | 0.891 |
|  | SqueezeBERT | 0.970 | 0.972 | 0.970 | 0.971 | 0.868 |

### Models performance

BERT, RoBERTa, DistilBERT, and SqueezeBERT all performed well after fine tuning, with BERT and RoBERTa performing the best with an F1-score of 0.976. Their macro F1-score were 0.865 and 0.891, respectively. Precision and recall of each class are presented for all models in Table S2. DistilBERT performed poorly in zero-shot classification, while DeBERTa’s performance seemed to scale with the number of parameters, reaching performance close to that of the smaller fine-tuned models for the large DeBERTa model with 304M parameters. Confusion matrices - calculated for the same test set for all the models - are available as Supplementary Materials in Figures S1 and S2. After analyzing the content of the incorrectly predicted classes, the main source of errors was situations where a weight loss or weight gain was mentioned in the text but did not refer to the drug covered by the comment. All comments affected by prediction errors contained syntax elements mentioning weight gain or loss, changes in appetite, as well as nausea and vomiting. Prediction errors were mainly due to a lack of attention to temporality in the narrative, to the mention of weight without any indication of change, or to the expression of a desire to gain or lose weight, whether or not related to the medication. In addition, it’s important to note that some prediction errors can be considered a posteriori as labeling errors. All of these misclassifications can be considered as false positives, indicating that the sensitivity of detecting changes in body weight would have been higher if we didn’t take into account the fact that the AE must be related to the drug at the source of the comment.

## Discussion

In this work, we explored the potential of large language models (LLMs) for detecting signals of body weight change as a side effects of antidepressants.

An important key point of this work is that we have built a valuable human annotated database, controlled by experts, with a substantial size of 8,000 input texts to evaluate model performance. We have made our dataset publicly available to promote open science and enable other researchers to build upon our labeling efforts (*doi to be released*). Collecting data on health-focused sites was relevant because it is estimated that only 10% of medical content on general social networks includes information on AEs, compared to 20-25% on health-focused platforms all AEs combined (19). Focusing on a single AE, we observed a proportion of 10% of the reviews made on antidepressant drugs were mentioning weight-related Aes - confirming that this AE might be a concern for patients taking these treatments. This high rate of AEs also confirmed that the choice of data source is important to reduce noise and identify AEs more easily.

In terms of model performance, the fine-tuned models were able to automatically distinguish situations of weight gain or loss associated with antidepressant treatment in user-generated online content. The performance of all the fine-tuned models were good - even for LLMs with a more limited number of parameters. We showed that LLMs can enable the detection of subtle signals within tens of thousands of comments pertaining to antidepressants, which could streamline the signal screening process. Zero-shot classification results are also very encouraging - especially with the large DeBERTa model. It has been demonstrated that pre-trained models with larger number of parameters shows better generalization results (44–46). It will be interesting to explore models with even more scaled properties in future works. Zero-shot approaches eliminate the need for annotation, making it possible to study a wide range of adverse events directly from extracted user reviews.

These models also tended to produce false positives. This is partly due to the annotation strategy, which focused only on mentions of body weight changes for the molecule being discussed in the review. Weight loss seems more difficult to classify and can be confused with weight gain - perhaps because these two classes share very similar syntactic elements. If an expert review of identified signals will likely remain essential for assessing a drug’s causality in the occurrence of adverse effects, it will be interesting to investigate how these models can aid in contextualizing the findings to support causality determination (47). Because online comments are often written with little contextual information, it is difficult to establish a causal relationship between drug use and the occurrence of an event. This might be the strongest limitation for the use of these data for pharmacovigilance.

Finally, extracting relevant medical information from online data depends on the quality of the data source. The ability of malicious social media bots to generate realistic comments raises doubts about the authenticity of all these comments and reviews, making it challenging to distinguish genuine patient feedback. (48–50). For this reason, the analysis of data from web platforms must be a matter of careful interpretation. Some specialized tools to detect fake reviews are probably expected as a safeguard. And if we can hope that website policies can partially prevent fake declarative comments and validate the credibility of their data, LLMs could be used at scale to provide insights for a better evaluation of medications in real-world conditions of use.

## Data Availability

Codes to reproduce tables and figures S1 and S2 are available in this public repository (git address to be specified)

## Funding information

This study did not receive any funding.

## Data and Code availability

Labeled data are available as an open source dataset on FigShare (DOI to be specified). Codes to reproduce tables and figures S1 and S2 are available in this public repository (git address to be specified)

## ACKNOWLEDGEMENTS

We acknowledge Maxime Alter, Furkan Erol, Ahmed Guendouz, Vitoria Morais-Brazil, Paule Nkeng, Inas Oulkaid-Mouaddan, Sofia Salaa and Chinar Salmanli for their valuable help in data annotation.

## Supplementary Note 1

**Table S1:** Distribution of the labels of the input texts for the different drugs in the whole dataset (n = 8,000)

| DRUG | No mention to weight change | Gain | Loss |
| --- | --- | --- | --- |
| Amitriptyline | 320 | 51 | 3 |
| Bupropion | 606 | 15 | 58 |
| Citalopram | 564 | 39 | 11 |
| Clomipramine | 49 | 5 | 1 |
| Desipramine | 8 | 1 | 0 |
| Desvenlafaxine | 228 | 8 | 11 |
| Doxepin | 84 | 19 | 0 |
| Duloxetine | 696 | 44 | 15 |
| Escitalopram | 863 | 96 | 22 |
| Esketamine | 10 | 0 | 0 |
| Fluoxetine | 455 | 23 | 15 |
| Fluvoxamine | 101 | 11 | 1 |
| Imipramine | 42 | 3 | 4 |
| Isocarboxazid | 1 | 3 | 0 |
| Levomilnacipran | 33 | 1 | 1 |
| Maprotiline | 3 | 0 | 0 |
| Milnacipran | 92 | 3 | 4 |
| Mirtazapine | 244 | 74 | 4 |
| Nefazodone | 43 | 0 | 0 |
| Nortriptyline | 136 | 21 | 2 |
| Paroxetine | 284 | 50 | 7 |
| Phenelzine | 28 | 6 | 1 |
| Protriptyline | 1 | 0 | 0 |
| Selegiline | 14 | 0 | 0 |
| Sertraline | 722 | 72 | 20 |
| Tranylcypromine | 21 | 2 | 1 |
| Trazodone | 409 | 6 | 1 |
| Trimipramine | 3 | 0 | 0 |
| Venlafaxine | 814 | 51 | 15 |
| Vilazodone | 177 | 9 | 4 |
| Vortioxetine | 123 | 8 | 4 |

## Supplementary Note 2

**Table S2:**
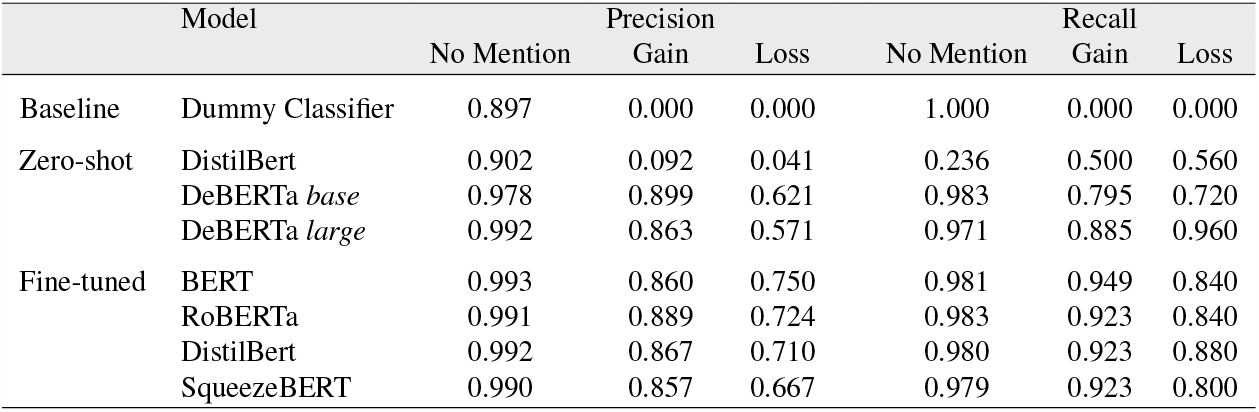
Per class precision and recall of the different BERT-based LLMs evaluated on a validation dataset of 1,000 text inputs

## Supplementary Note 3

**Fig S1:**
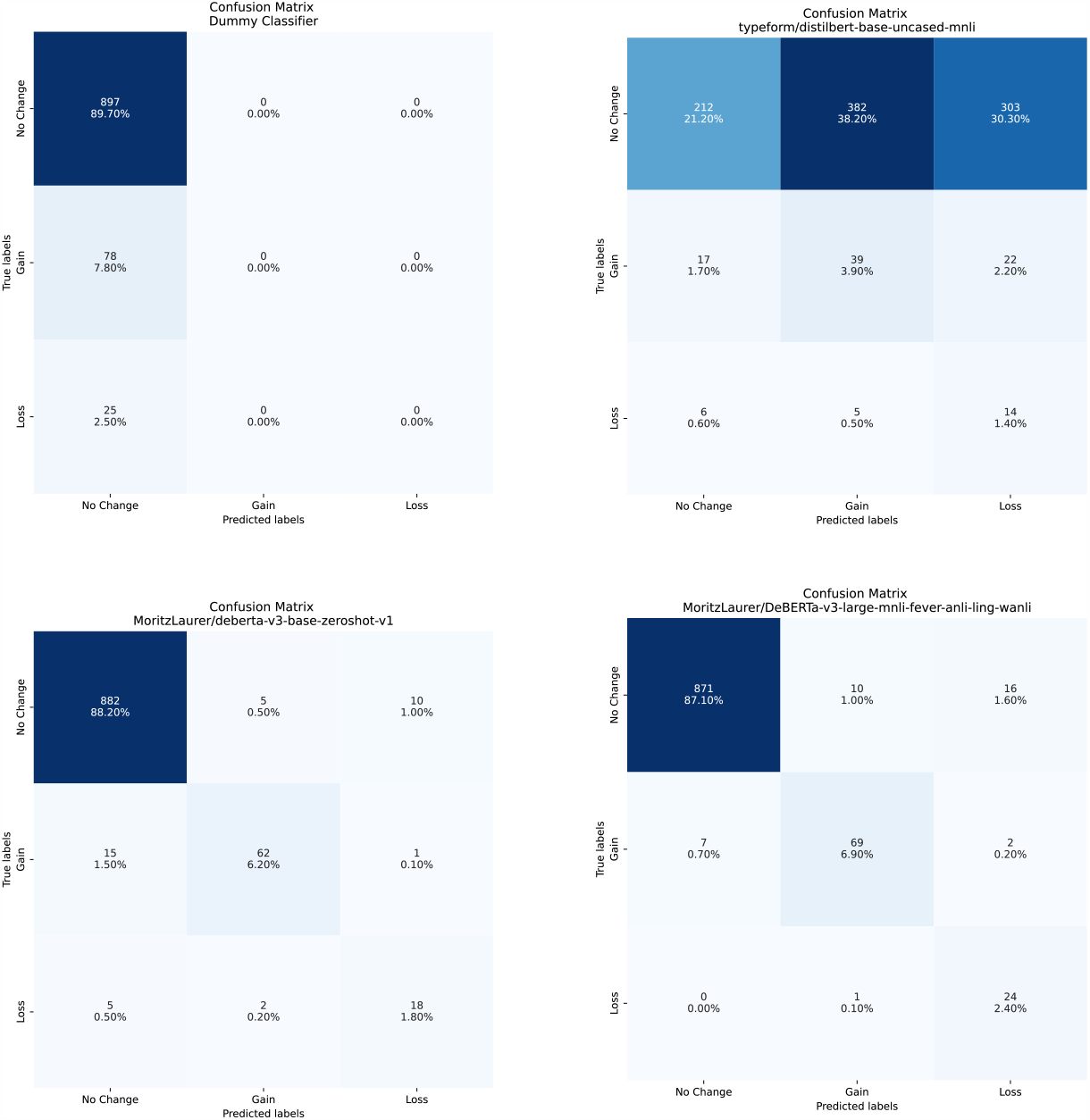
Confusion matrices of the baseline and zero-shot models on the target domain, showing the predicted vs. true labels. Performance metrics of the models are described in Table 1 and Table S2

## Supplementary Note 4

**Fig S2:**
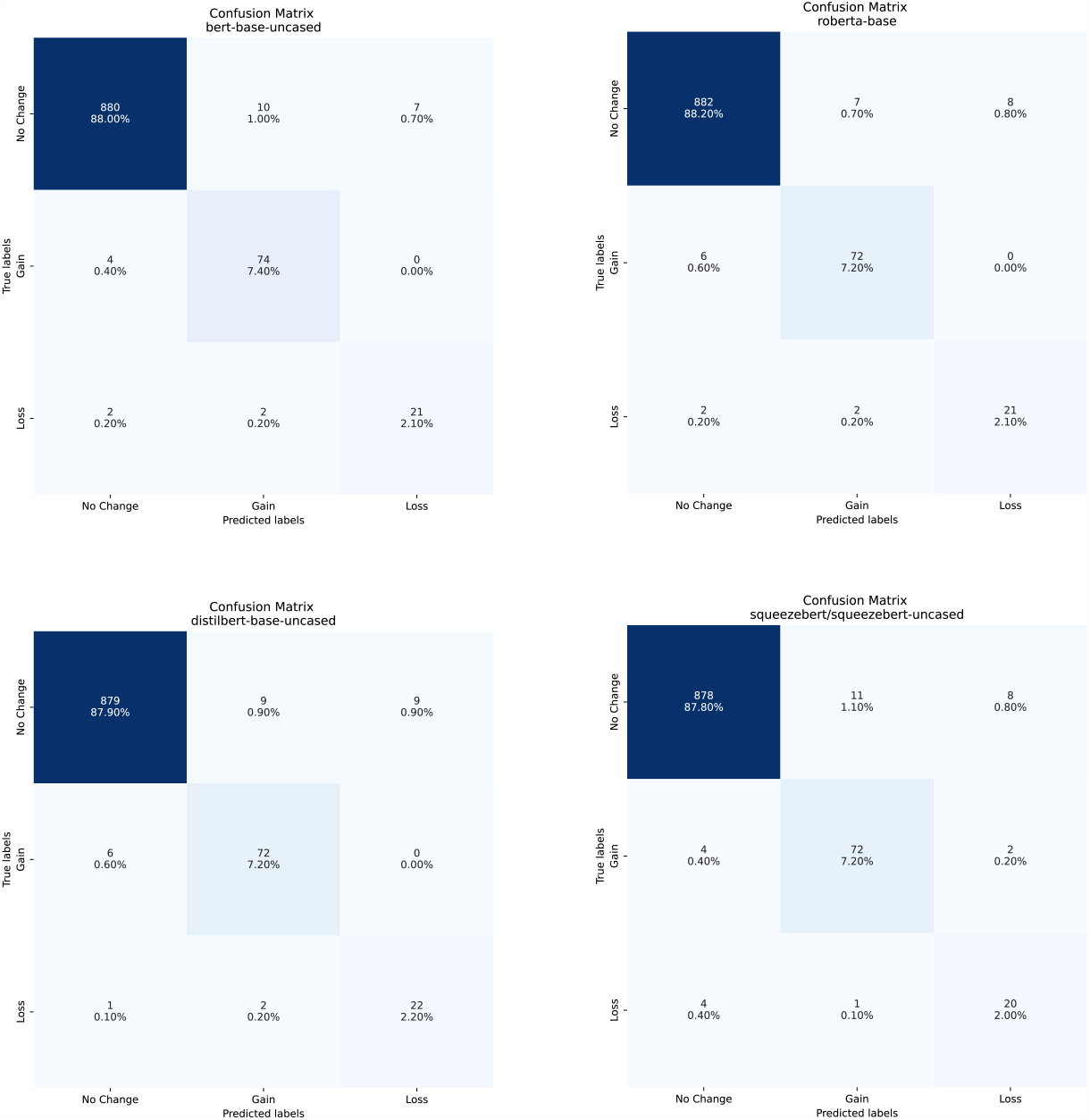
Confusion matrices of the fine-tuned models on the target domain, showing the predicted vs. true labels. Performance metrics of the models are described in Table 1 and Table S2

## Notes

### Competing Interest Statement

The authors have declared no competing interest.

## Bibliography

1. Alessandro Serretti and Laura Mandelli. Antidepressants and body weight: A comprehensive review and meta-analysis. Journal of Clinical Psychiatry, 71(10):1259–1272, oct 2010. ISSN 01606689. doi: 10.4088/JCP.09r05346blu.

2. Gilbert H Berken, Dorothy O Weinstein, and Warren C Stern. Weight gain: a side-effect of tricyclic antidepressants. Journal of Affective Disorders, 7(2):133–138, 1984.

3. Juan Pablo Domecq, Gabriela Prutsky, Aaron Leppin, M. Bassam Sonbol, Osama Altayar, Chaitanya Undavalli, Zhen Wang, Tarig Elraiyah, Juan Pablo Brito, Karen F. Mauck, Mohammed H. Lababidi, Larry J. Prokop, Noor Asi, Justin Wei, Salman Fidahussein, Victor M. Montori, and Mohammad Hassan Murad. Drugs commonly associated with weight change: A systematic review and meta-analysis. Journal of Clinical Endocrinology and Metabolism, 100(2):363–370, 2015. ISSN 19457197. doi: 10.1210/jc.2014-3421.

4. Rafael Gafoor, Helen P. Booth, and Martin C. Gulliford. Antidepressant utilisation and incidence of weight gain during 10 years’ follow-up: Population based cohort study. BMJ (Online), 361, may 2018. ISSN 17561833. doi: 10.1136/bmj.k1951.

5. Lauren B. Solberg. The benefits of online health communities. Virtual Mentor, 16(4):270–274, apr 2014. ISSN 19377010. doi: 10.1001/virtualmentor.2014.16.4.stas1-1404.

6. Marianne Wikgren. Everyday health information exchange and citation behaviour in Internet discussion groups. The New Review of Information Behaviour Research, 4(1):225–239, 2003. ISSN 1471-6313. doi: 10.1080/14716310310001631543.

7. Yasser M. Alatawi and Richard A. Hansen. Empirical estimation of under-reporting in the U.S. Food and Drug Administration Adverse Event Reporting System (FAERS). Expert Opinion on Drug Safety, 16(7):761–767, 2017. ISSN 1744764X. doi: 10.1080/14740338.2017.1323867.

8. Lorna Hazell and Saad A.W. Shakir. Under-reporting of adverse drug reactions: A systematic review. Drug Safety, 29(5):385–396, 2006. ISSN 01145916. doi: 10.2165/00002018-200629050-00003.

9. Diksha Khurana, Aditya Koli, Kiran Khatter, and Sukhdev Singh. Natural language processing: state of the art, current trends and challenges. Multimedia Tools and Applications, 82 (3):3713–3744, 2023. ISSN 15737721. doi: 10.1007/s11042-022-13428-4.

10. Diego Saldana Miranda. Automated detection of adverse drug reactions in the biomedical literature using convolutional neural networks and biomedical word Embeddings. CEUR Workshop Proceedings, 2226:33–41, 2018. ISSN 16130073.

11. Elena Tutubalina and Sergey Nikolenko. Combination of Deep Recurrent Neural Networks and Conditional Random Fields for Extracting Adverse Drug Reactions from User Reviews. Journal of Healthcare Engineering, 2017, 2017. ISSN 20402309. doi: 10.1155/2017/9451342.

12. Min Zhang and Guohua Geng. Adverse drug event detection using a weakly supervised convolutional neural network and recurrent neural network model. Information (Switzerland), 10(9):276, 2019. ISSN 20782489. doi: 10.3390/info10090276.

13. B. V. Anjali and G. K. Ravi Kumar. A Broad Review on Adverse Drug Reaction Detection using Social Media Data. In Proceedings - 2022 6th International Conference on Intelligent Computing and Control Systems, ICICCS 2022, pages 1852–1856. IEEE, 2022. ISBN 9781665410359. doi: 10.1109/ICICCS53718.2022.9788381.

14. Yuan Luo, William K. Thompson, Timothy M. Herr, Zexian Zeng, Mark A. Berendsen, Siddhartha R. Jonnalagadda, Matthew B. Carson, and Justin Starren. Natural Language Processing for EHR-Based Pharmacovigilance: A Structured Review. Drug Safety, 40(11): 1075–1089, 2017. ISSN 11791942. doi: 10.1007/s40264-017-0558-6.

15. Susmitha Wunnava, Xiao Qin, Tabassum Kakar, Cansu Sen, Elke A. Rundensteiner, and Xiangnan Kong. Adverse Drug Event Detection from Electronic Health Records Using Hierarchical Recurrent Neural Networks with Dual-Level Embedding. Drug Safety, 42(1):113–122, 2019. ISSN 11791942. doi: 10.1007/s40264-018-0765-9.

16. Heba Ibrahim, A. Abdo, Ahmed M. El Kerdawy, and A. Sharaf Eldin. Signal Detection in Pharmacovigilance: A Review of Informatics-driven Approaches for the Discovery of Drug-Drug Interaction Signals in Different Data Sources. Artificial Intelligence in the Life Sciences, 1:100005, 2021. ISSN 26673185. doi: 10.1016/j.ailsci.2021.100005.

17. Rachel M. Murphy, Joanna E. Klopotowska, Nicolette F. de Keizer, Kitty J. Jager, Jan Hendrik Leopold, Dave A. Dongelmans, Ameen Abu-Hanna, and Martijn C. Schut. Adverse drug event detection using natural language processing: A scoping review of supervised learning methods. PLoS ONE, 18(1 January), 2023. ISSN 19326203. doi: 10.1371/journal.pone.0279842.

18. Robert Leaman, Laura Wojtulewicz, Ryan Sullivan, Annie Skariah, Jian Yang, and Graciela Gonzalez. Towards internet-age pharmacovigilance: Extracting adverse drug reactions from user posts to health-related social networks. In Proceedings of the Annual Meeting of the Association for Computational Linguistics, pages 117–125, 2010. ISBN 1932432736.

19. Abeed Sarker, Rachel Ginn, Azadeh Nikfarjam, Karen O’Connor, Karen Smith, Swetha Jayaraman, Tejaswi Upadhaya, and Graciela Gonzalez. Utilizing social media data for pharmacovigilance: A review. Journal of Biomedical Informatics, 54:202–212, 2015. ISSN 15320464. doi: 10.1016/j.jbi.2015.02.004.

20. Azadeh Nikfarjam, Julia D. Ransohoff, Alison Callahan, Erik Jones, Brian Loew, Bernice Y. Kwong, Kavita Y. Sarin, and Nigam H. Shah. Early detection of adverse drug reactions in social health networks: A natural language processing pipeline for signal detection. JMIR Public Health and Surveillance, 5(2), 2019. ISSN 23692960. doi: 10.2196/11264.

21. Yefeng Wang, Yunpeng Zhao, Dalton Schutte, Jiang Bian, and Rui Zhang. Deep learning models in detection of dietary supplement adverse event signals from Twitter. JAMIA Open, 4(4), 2021. ISSN 25742531. doi: 10.1093/jamiaopen/ooab081.

22. Arjun Magge, Abeed Sarker, Azadeh Nikfarjam, and Graciela Gonzalez-Hernandez. Comment on: “Deep learning for pharmacovigilance: Recurrent neural network architectures for labeling adverse drug reactions in Twitter posts”. Journal of the American Medical Informatics Association, 26(6):577–579, 2019. ISSN 1527974X. doi: 10.1093/jamia/ocz013.

23. Ashish Vaswani, Noam Shazeer, Niki Parmar, Jakob Uszkoreit, Llion Jones, Aidan N. Gomez, Łukasz Kaiser, and Illia Polosukhin. Attention is all you need. Advances in Neural Information Processing Systems, 2017-December:5999–6009, 2017. ISSN 10495258.

24. Benjamin Kompa, Joe B. Hakim, Anil Palepu, Kathryn Grace Kompa, Michael Smith, Paul A. Bain, Stephen Woloszynek, Jeffery L. Painter, Andrew Bate, and Andrew L. Beam. Correction to: Artificial Intelligence Based on Machine Learning in Pharmacovigilance: A Scoping Review (Drug Safety, (2022), 45, 5, (477-491), 10.1007/s40264-022-01176-1). Drug Safety, 46(4):433, 2023. ISSN 11791942. doi: 10.1007/s40264-023-01273-9.

25. Amy Breden and Lee Moore. Detecting Adverse Drug Reactions from Twitter through Domain-Specific Preprocessing and BERT Ensembling. arXiv preprint arXiv:2005.06634, 2020.

26. George Andrei Dima, Dumitru Clementin Cercel, and Mihai Dascalu. Transformer-based Multi-Task Learning for Adverse Effect Mention Analysis in Tweets. In Social Media Mining for Health, SMM4H 2021 - Proceedings of the 6th Workshop and Shared Tasks, pages 44–51, 2021. ISBN 9781954085312. doi: 10.18653/v1/2021.smm4h-1.7.

27. Sidharth Ramesh, Abhiraj Tiwari, Parthivi Choubey, Saisha Kashyap, Sahil Khose, Kumud Lakara, Nishesh Singh, and Ujjwal Verma. BERT based Transformers lead the way in Extraction of Health Information from Social Media. In Social Media Mining for Health, SMM4H 2021 - Proceedings of the 6th Workshop and Shared Tasks, pages 33–38, 2021. ISBN 9781954085312. doi: 10.18653/v1/2021.smm4h-1.5.

28. Simone Scaboro, Beatrice Portelli, Emmanuele Chersoni, Enrico Santus, and Giuseppe Serra. Extensive evaluation of transformer-based architectures for adverse drug events extraction. Knowledge-Based Systems, 275:110675, 2023. ISSN 09507051. doi: 10.1016/j.knosys.2023.110675.

29. Yu Gu, Sheng Zhang, Naoto Usuyama, Yonas Woldesenbet, Cliff Wong, Praneeth Sanapathi, Mu Wei, Naveen Valluri, Erika Strandberg, Tristan Naumann, and Hoifung Poon. Distilling Large Language Models for Biomedical Knowledge Extraction: A Case Study on Adverse Drug Events. arXiv preprint arXiv:2307.06439, 2023.

30. Wenpeng Yin, Jamaal Hay, and Dan Roth. Benchmarking zero-shot text classification: Datasets, evaluation and entailment approach. EMNLP-IJCNLP 2019 - 2019 Conference on Empirical Methods in Natural Language Processing and 9th International Joint Conference on Natural Language Processing, Proceedings of the Conference, pages 3914–3923, 2019. doi: 10.18653/v1/d19-1404.

31. Chi Sun, Xipeng Qiu, Yige Xu, and Xuanjing Huang. How to Fine-Tune BERT for Text Classification? In Lecture Notes in Computer Science (including subseries Lecture Notes in Artificial Intelligence and Lecture Notes in Bioinformatics), volume 11856 LNAI, pages 194–206. Springer, 2019. ISBN 9783030323806. doi: 10.1007/978-3-030-32381-3_16.

32. Jacob Devlin, Ming Wei Chang, Kenton Lee, and Kristina Toutanova. BERT: Pre-training of deep bidirectional transformers for language understanding. NAACL HLT 2019 - 2019 Conference of the North American Chapter of the Association for Computational Linguistics: Human Language Technologies - Proceedings of the Conference, 1:4171–4186, 2019.

33. Yinhan Liu, Myle Ott, Naman Goyal, Jingfei Du, Mandar Joshi, Danqi Chen, Omer Levy, Mike Lewis, Luke Zettlemoyer, and Veselin Stoyanov. RoBERTa: A Robustly Optimized BERT Pretraining Approach. arXiv preprint arXiv:1907.11692, 2019.

34. Victor Sanh, Lysandre Debut, Julien Chaumond, and Thomas Wolf. DistilBERT, a distilled version of BERT: smaller, faster, cheaper and lighter. arXiv preprint arXiv:1910.01108, 2019.

35. Forrest Iandola, Albert Shaw, Ravi Krishna, and Kurt Keutzer. SqueezeBERT: What can computer vision teach NLP about efficient neural networks? arXiv preprint arXiv:2006.11316, pages 124–135, 2020. doi: 10.18653/v1/2020.sustainlp-1.17.

36. Yi Tay, Mostafa Dehghani, Jinfeng Rao, William Fedus, Samira Abnar, Hyung Won Chung, Sharan Narang, Dani Yogatama, Ashish Vaswani, and Donald Metzler. Scale efficiently: Insights from pre-training and fine-tuning transformers. arXiv preprint arXiv:2109.10686, 2021.

37. Pengcheng He, Xiaodong Liu, Jianfeng Gao, and Weizhu Chen. Deberta: Decoding-Enhanced Bert With Disentangled Attention. ICLR 2021 - 9th International Conference on Learning Representations, 2021.

38. Joshua Hardwick. Top 100 most visited websites in the world (as of 2020), 2020.

39. Hadley Wickham. rvest: Easily Harvest (Scrape) Web Pages. R package version 0.3.1. https://CRAN.R-project.org/package=rvest, 2015.

40. Hadley Wickham. httr: Tools for Working with URLs and HTTP. R package version 1.1.0., 2016.

41. Wickham Hadley, Hester Jim, and Ooms Jeroen. xml2: Parse XML. R package, 2021.

42. Thomas Wolf, Lysandre Debut, Victor Sanh, Julien Chaumond, Clement Delangue, Anthony Moi, Pierric Cistac, Tim Rault, Rémi Louf, Morgan Funtowicz, and Others. Transformers: State-of-the-art natural language processing. In Proceedings of the 2020 conference on empirical methods in natural language processing: system demonstrations, pages 38–45, 2020.

43. Ilya Loshchilov and Frank Hutter. Decoupled weight decay regularization. 7th International Conference on Learning Representations, ICLR 2019, 2019.

44. Colin Raffel, Noam Shazeer, Adam Roberts, Katherine Lee, Sharan Narang, Michael Matena, Yanqi Zhou, Wei Li, and Peter J. Liu. Exploring the limits of transfer learning with a unified text-to-text transformer. Journal of Machine Learning Research, 21(140):1–67, 2020. ISSN 15337928.

45. Tom B. Brown, Benjamin Mann, Nick Ryder, Melanie Subbiah, Jared Kaplan, Prafulla Dhariwal, Arvind Neelakantan, Pranav Shyam, Girish Sastry, Amanda Askell, Sandhini Agarwal, Ariel Herbert-Voss, Gretchen Krueger, Tom Henighan, Rewon Child, Aditya Ramesh, Daniel M. Ziegler, Jeffrey Wu, Clemens Winter, Christopher Hesse, Mark Chen, Eric Sigler, Mateusz Litwin, Scott Gray, Benjamin Chess, Jack Clark, Christopher Berner, Sam McCandlish, Alec Radford, Ilya Sutskever, and Dario Amodei. Language models are few-shot learners. Advances in Neural Information Processing Systems, 2020-December:1877–1901, 2020. ISSN 10495258.

46. Mohammad Shoeybi, Mostofa Patwary, Raul Puri, Patrick LeGresley, Jared Casper, and Bryan Catanzaro. Megatron-LM: Training Multi-Billion Parameter Language Models Using Model Parallelism. arXiv preprint arXiv:1909.08053, 2019.

47. Sajid Hussain, Hammad Afzal, Ramsha Saeed, Naima Iltaf, and Mir Yasir Umair. Pharmacovigilance with Transformers: A Framework to Detect Adverse Drug Reactions Using BERT Fine-Tuned with FARM. Computational and Mathematical Methods in Medicine, 2021, 2021. ISSN 17486718. doi: 10.1155/2021/5589829.

48. Mariam Orabi, Djedjiga Mouheb, Zaher Al Aghbari, and Ibrahim Kamel. Detection of Bots in Social Media: A Systematic Review. Information Processing and Management, 57(4):102250, 2020. ISSN 03064573. doi: 10.1016/j.ipm.2020.102250.

49. Maxim Kolomeets and Andrey Chechulin. HadleyAnalysis of the Malicious Bots Market. In Conference of Open Innovation Association, FRUCT, volume 2021-May, pages 199–205. IEEE, 2021. ISBN 9789526924458. doi: 10.23919/FRUCT52173.2021.9435421.

50. Anahita Davoudi, Ari Z Klein, Abeed Sarker, and Graciela Gonzalez-Hernandez. HadleyTowards Automatic Bot Detection in Twitter for Health-related Tasks. AMIA Joint Summits on Translational Science proceedings. AMIA Joint Summits on Translational Science, 2020:136–141, 2020. ISSN 2153-4063.

